# Psychometric Properties of the NIH Toolbox Cognition and Emotion Batteries Among Children and Adolescents with Congenital Heart Defects

**DOI:** 10.1101/2023.02.11.23285800

**Authors:** Julia Wallace, Rafael Ceschin, Vince K. Lee, Nancy H. Beluk, Cheryl Burns, Sue Beers, Cecilia Lo, Ashok Panigrahy, Daryaneh Badaly

## Abstract

**Objective:** The NIH Toolbox offers brief, computerized measures of cognitive and psychosocial functioning. However, its psychometric properties were established among typically developing children and adolescents. The current study provides the first comprehensive assessment of its psychometric properties among young patients with congenital heart defects (CHD).

**Study Design:** We prospectively recruited 58 patients with CHD and 80 healthy controls between the ages of 6 and 17. Participants completed the NIH Toolbox Cognition and Emotion Batteries, a battery of clinician-administered neuropsychological tests, and ratings of their quality of life. Their parents also completed ratings of their functioning.

**Results:** On the Cognition Battery, we found expectable group differences and developmentally expected gains across ages. For the most part, composites and subtests were significantly correlated with neuropsychological measures of similar constructs. Higher scores were generally associated with ratings of better day-to-day functioning among children with CHD. On the Emotion Battery, we found no significant group differences, echoing prior research. For the most part, scales showed acceptable internal consistency among both groups. There was adequate construct coherence for most of questionnaires among healthy control but not participants with CHD. Correlations with a comparison tool were largely within expectable directions.

**Conclusion:** The NIH Toolbox may provide a valid and useful assessment of cognitive functioning among children and adolescents with CHD. While it may offer reliable and valid scales of psychosocial functioning, further research is needed to understand the meaningfulness of the scales for participants with CHD.

---

Children and adolescents with congenital heart defects (CHD), particularly those with more severe forms of cardiac anomalies, those who have undergone surgical correction in the first year of life, and those with medical comorbidities, are at risk for a myriad of cognitive and psychosocial challenges (Phillips & Longoria, 2020). As a result, guidelines and recommendations have been set forth for the screening, assessment, and intervention of neurodevelopmental concerns among young individuals with CHD (Marino et al., 2012; Ilardi et al., 2020). With this in mind, the development of a uniform set of brief and easily administered assessment tools may facilitate screening, tracking progress over time, and comparing data across centers. The cognitive and psychosocial assessments from the NIH Toolbox (NIHTB) may offer such a set of tools. However, their psychometric properties have yet to be explored among children and adolescents with CHD.

The NIHTB offers a series of computerized assessments of different domains of functioning (www.nihtoolbox.org; Gershon et al., 2013). In particular, the NIHTB Cognition Battery (NIHTB-CB) and Emotion Battery (NIHTB-EB) assess cognitive and psychosocial functioning. The NIHTB was designed to provide a set of common data elements among disparate centers using standard methodology to minimize the likelihood that result differences would be attributable to the test instruments used. It was normed for ages 3 to 85 providing a set of tools to study outcomes across the lifespan, and has been translated into a number of languages to allow for global comparisons. Furthermore, the NIHTB is both user- and participant-friendly, as it provides accessible and flexible training options, can be completed in relatively brief amount of time by participants via in person or virtual administration, and offers automatic calculation of scores.

The psychometric properties of the NIHTB were established among typically developing individuals (Bauer & Zelazo, 2013; Mungas et al., 2013; Salsman et al., 2013). However, its authors highlighted the importance of validation among clinical populations (Weintraub et al., 2013). Recently, researchers have begun to use the NIHTB among individuals with CHD, a population with an increased risk of deficits across multiple cognitive and socioemotional domains (Bellinger & Newburger, 2013). That being said, the psychometric properties of the tool within this population have largely remained unexplored. As a result, the current study examined the psychometric properties NIHTB-CB and NIHTB-EB among children and adolescents with CHD, as compared to healthy controls. We examined whether the tools showed: a) expectable group similarities and differences among patients with CHD and healthy controls, b) typical developmental trends (for cognitive tasks only), c) coherent structures using confirmatory factor analysis and indices of internal consistency (for psychosocial questionnaires only), d) correlations with measures of similar constructs (i.e., convergent validity), and d) and anticipated correlations with measures of day-to-day functioning (i.e., concurrent validity) (for cognitive tasks only).

## Methods

### Participants

We prospectively recruited 97 children and adolescents with an array of heart defects, including those with single ventricle physiology, aortic arch anomalies, stenosis, and other malformations, as well as 88 healthy controls from a single academic medical center. We recruited English-speaking children and adolescents between 6 to 17 years of age. Individuals diagnosed with chromosomal anomalies or with history of intensive treatment for any diagnosis were excluded from the control group. Of the 185 recruited participants for whom parents provided consent, 43 individuals were not assessed due to the following reasons: scheduling conflicts (26), decided to longer participate or withdrew (10), behavioral noncompliance (4), or other reasons (3). Additionally, 3 healthy participants with significant medical histories and 1 participant with CHD who underwent a heart transplant after consent were withdrawn by the principal investigators. The final sample of 138 participants included 58 subjects with CHD and 80 healthy controls. Details of the demographics and diagnoses of the participants are summarized in Supplemental Table 1.

Children were assented to the project, and their parent or legal guardian provided consent on their behalf. The project was approved by the University of Pittsburgh Institutional Review Board and completed in accordance with the ethical principles of the Helsinki Declaration.

### Assessment Instruments

Fifty-eight participants with CHD and 74 comparison controls completed the NIHTB-CB using both desktop and iPad versions of the battery; 52 children and adolescents with CHD and 68 healthy controls responded to self-report questionnaires from the NIHTB-EB. A clinician-issued pencil-and-paper battery was administered to a subset of 44 participants with CHD and 70 controls; parents and participants also completed behavioral ratings and quality of life inventories.

#### NIHTB Cognitive Battery

Participants completed subtests of NIHTB-CB, generating composite scores for Crystallized Cognition, Fluid Cognition, and Total Cognition. The subtests for Crystallized Cognition include the Oral Reading Recognition Test (ORRT) and the Picture Vocabulary Test (PVT). On the ORRT, a test of letter identification and word reading, participants were shown letters (for younger children) or words (for older participants) and asked to read them aloud (Gershon et al., 2013). On the PVT, which assesses receptive vocabulary, participants selected images matching descriptive audio cues. The test uses computer adaptive testing methods (Gershon et al., 2013).

The subtests for Fluid Cognition include the List Sorting Working Memory Test (LSWMT), the Pattern Comparison Processing Speed Test (PCPST), the Flanker Inhibitory Control and Attention Test (FIC+AT), the Dimensional Change Card Sort Test – Executive Function (DCCST), and the Picture Sequence Memory Test (PSMT). On the LSWMT, a test of working memory, participants ages 7 and older were presented with a series of items presented visually and auditorily and were then asked to repeat the presented items, ordering them based on particular criteria (e.g., item size). The test requires participants to hold a set amount of information in mind, mentally organize the list of items based on a given criteria, and then verbally recall the information (Tulsky et al., 2013). On the PCPST, a test of processing speed, participants were presented with two images on the screen and had to quickly indicate if the two images matched (smiley and frowny face buttons for those ages 6 and younger, yes and no buttons for those ages 7 and older) (Carlozzi et al., 2013). On the FIC+AT, participants were instructed to focus on a central image (fish for those ages 7 and younger; arrows for those ages 8 and older) flanked by additional stimuli, which may or may not point in the same direction. Participants then had to identify the direction of the middle stimulus. The test assesses the ability to selectively pay attention to stimuli while inhibiting focus on irrelevant information (Zelazo et al., 2013). On the DCCST, which requires inhibitory control and mental flexibility, participants were first instructed to focus on the color or shape of a forthcoming image and then shown a reference image. Depending on the prompt, participants had to quickly match one of two test images, varying in both color and shape, to the reference image (Zelazo et al., 2013). On the PSMT, participants were presented with a series of pictorial scenes presented in a particular order, and then had to identify the order in which the scenes were presented. The test evaluates learning and immediate retrieval of information (Bauer et al., 2013). For each subtest of the NIHTB-CB, we derived age-corrected standard scores (*M* = 100, *SD* = 15).

#### NIHTB Emotion Battery

The NIHTB-EB includes measures of negative affect, psychosocial well-being, stress and self-efficacy, and social relationships (Salsman et al., 2013). Specifically, there are three scales assessing negative affect or experiences of unpleasant or distressing emotions (i.e., Anger, Fear, and Sadness), two scales assessing psychological well-being or feelings of pleasure and contentment (i.e., Positive Affect and General Life Satisfaction), two scales assessing stress and self-efficacy or one’s perception of everyday experiences and ability to respond to challenging events (i.e., Perceived Stress and Self-Efficacy), and five scales assessing social relationships, with a focus on one’s perception of the availability and quality of those relationships (i.e., Emotional Support, Loneliness, Friendship, Perceived Hostility, and Perceived Rejection). Each of the scales of the NIHTB-EB is completed on a 5-point Likert scale. While some of the scales assess 8- to 17-year-olds, others have distinct forms for 8- to 12-year-olds and 13- to 17-year-olds, with developmentally appropriate wording and item banks. All of the scales for 8- to 12-year-olds and the majority for 13- to 17-year-olds used a fixed number of items; three scales for 13- to 17-year-olds (i.e., Positive Affect, General Life Satisfaction, and Self-Efficacy) used computer-adaptive testing (CAT) methods, in which items are selected from a bank based on participants’ progressive responses. For each scale, we derived fully-corrected *t*-scores (*M* = 50, *SD* = 10).

#### Clinician-Issued Battery and Rating Scales

For analyses examining convergent and concurrent validity with the NIHTB-CB, participants completed a clinician-issued battery of neuropsychological tests as part of a larger study. They completed the Wechsler Abbreviated Scale of Intelligence, 2^nd^ edition (WASI-II), a brief assessment of intellectual functioning comprised of four subtests, which provides composite scores of Verbal Reasoning, Fluid Reasoning, and Full Scale IQ. The Vocabulary subtest of the WASI-II assesses crystallized knowledge of vocabulary terms, and can be used in a similar manner as word reading tests to estimate functioning (Bright & van der Linde, 2020). To assess receptive language, participants up to age 16 completed the Comprehension of Instructions subtest from the Neuropsychology Assessment, 2^nd^ edition (NEPSY-2), in which they were asked to listen to oral instructions of increasing syntactic complexity and point to appropriate stimuli provided. Participants up to age 16 furthermore completed subtests from the Working Memory Index (Digit Span and Letter-Number Sequencing) and Processing Speed Index (Coding and Symbol Search) of the Wechsler Intelligence Scale for Children, 4^th^ edition (WISC-IV). Three tests of executive functioning from the Delis-Kaplan Executive Function Scale (D-KEFS) were administered to participants above age 8. Specifically, participants completed: the Color-Word Interference Test (CWIT), a variation of the classic Stroop test that requires inhibition of automatic responses when naming stimuli color or word; the Trail Making Test (TMT), which includes a trial of mental flexibility asking individuals to quickly switch between connecting circles containing numbers and letters in order; and, the Verbal Fluency Test (VFT), which includes a trial of mental flexibility asking individuals quickly switch between saying words in different categories. Lastly, participants completed the Design Memory subtest of the Wide Range Assessment of Memory and Learning, 2^nd^ edition (WRAML-2), a measure of visual learning and memory.

Participants and their parents filled out the Pediatric Quality of Life Inventory (PedsQL), rating scales measuring children’s overall adjustment as well as their adjustment in physical, emotional, school and social domains (with the later three also summarized to index psychosocial adjustment). Parents also completed ratings scales assessing children’s day-to-day adjustment across multiple domains. Parents completed the Adaptive Behavior Assessment System, 2^nd^ edition (ABAS-II), a questionnaire designed to provide estimates of adaptive behavior, including a measure of overall adaptive skill as well as performance in conceptual, social, and practical domains. Parent-ratings were also collected on the Behavior Assessment System for Children, 2^nd^ edition (BASC-2), which assesses multiple areas of emotional, behavioral, and adaptive functioning.

### Analysis Plan

We examined *group differences* between patients with CHD and healthy controls using independent sample *t*-tests for continuous variables and χ^2^-tests for frequencies within categorical variables. We considered differences in demographic variables, scores on the NIHTB, and scores on the clinician-issued battery and rating scales. We considered *developmental trends* for the NIHTB by analyzing the correlations between raw scores and age. We assessed whether measures on the NIHTB demonstrated adequate *convergent validity* by analyzing the correlations between test scores on the NIHTB and those on measures of similar abilities. We examined whether measures on the NIHTB demonstrated adequate *concurrent validity* by analyzing the correlations between test scores on the NIHTB and measures of day-to-day functioning. To understand if developmental trends or validity indices differed between participants with CHD and healthy controls, we used Fisher (1915; 1921)’s *r*-to-*z* transformation. To understand the *construct coherence* of questionnaires, we conducted confirmatory factor analyses (CFA). Because measures that were administered as computer adaptive tests did not have data for each of the questionnaire items, data were imputed using multiple imputation by chained equations (MICE) (Bulut & Kim, 2021). MICE was completed using an algorithm provided by the scikit-learn multiple imputation library (IterativeImputer) with a Bayesian ridge estimator and 500 iterations. Each participant’s response variables were used to estimate the missing data. A second analysis included both the response variables and group to estimate the missing data. Because the pattern of findings was similar with both imputations, only the latter findings are reported. We examined multiple measures of fit for our CFA models, including the standardized root mean square residual (SRMR), the root mean square error of approximation (RMSEA), and the Bentler comparative fit index (CFI). Adequate model fit was indicated by SRMR ≤ 0.12, RMSEA ≤ 0.12, and CFI ≥ 0.88, using guidelines from Taasoobshirazi & Wang, 2016. To explore the *internal consistency* of questionnaires, Cronbach’s alpha was calculated for measures that were administered as fixed forms, and marginal reliability coefficients were estimated with graded item response theory (IRT) models for measures that were administered as computer adaptive tests.

## Results

Participant demographics are detailed in Supplemental Table 1. There were larger percentages of individuals identified as White and as male among participants with CHD as compared to healthy controls. Otherwise, the two groups of participants did not differ on demographic variables.

### Cognitive Battery

#### Group Differences

Participants with CHD performed significantly poorer than healthy peers on the NIHTB-CB (*p*s ≤ 0.045), with exception of the LSWMT (Table 1). Still, children and adolescents with CHD generally performed within normal limits. Similarly, on the clinician-issued battery of cognitive tests, participants with CHD tended to perform more poorly than healthy controls but also tended to perform within normal limits (for the comparison data, see Supplemental Table 2). Of note, the overall cognitive level of healthy controls differed from the population mean based on both the NIHTB-CB Total Cognition (*t* = 5.78, *p* < 0.001) and the WASI-II Full Scale IQ (*t* = 6.26, *p* < 0.001). Similarly, the overall cognitive level of youths with CHD was higher than has been reported in past work (Feldman et al. 2021), suggesting that we captured typical group differences even if participants in both groups were slightly higher functioning than would be expected.

**Table 1.**
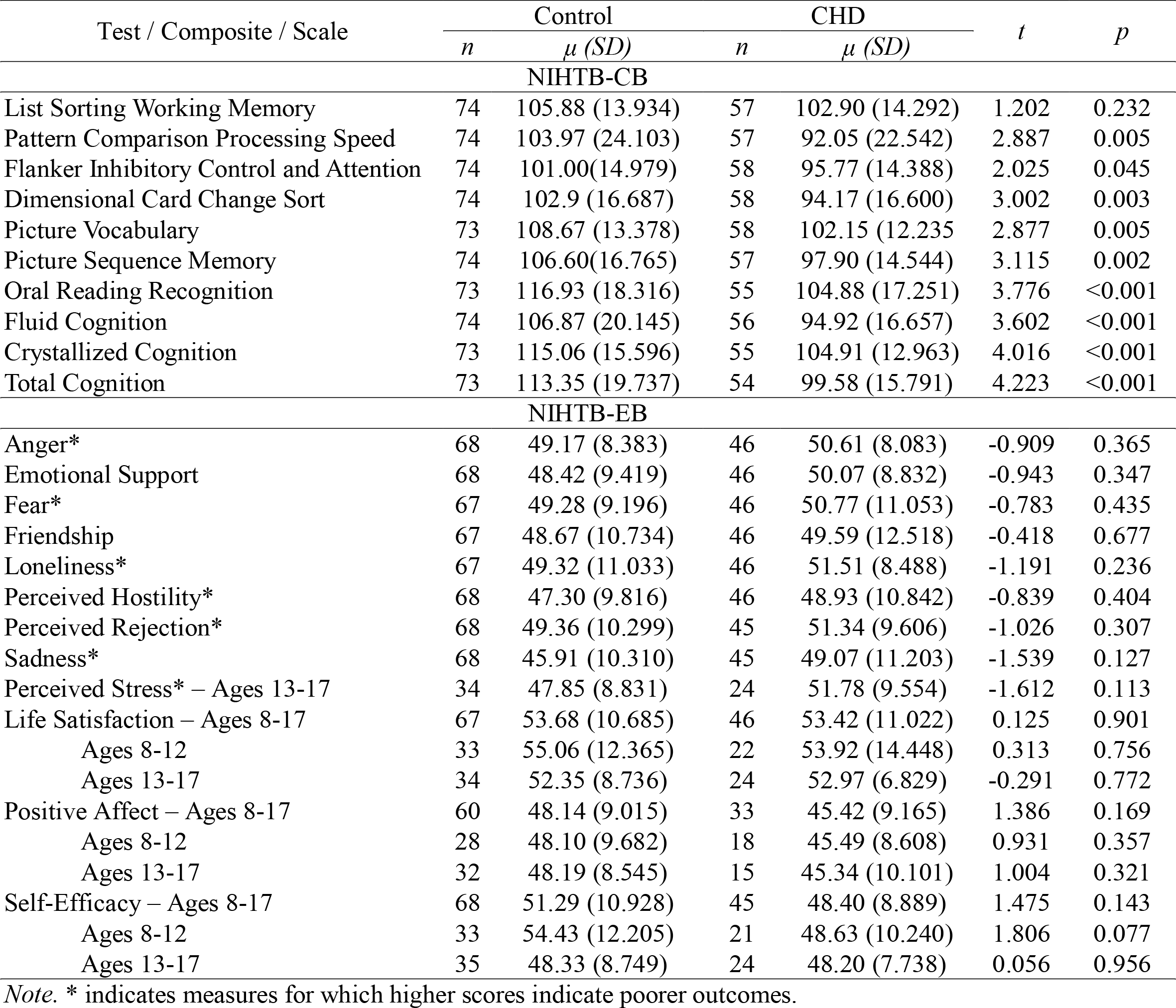
Overview of Group Differences: NIHTB-CB and NIHTB-EB

#### Developmental Trends

Age was positively associated with improved performance on all NIHTB-CB subtests and composite scores, both among participants with CHD (*rs* ≥ 0.547, *p*s < 0.001) and healthy controls (*r*s ≥ 0.400, *p*s < 0.001). Age effects did not significant differ between participants with CHD and healthy controls, based on Fisher’s r-to-z transformations (*z*s ≤ |1.71|, *p*s > 0.08).

#### Convergent Validity

As shown in Table 2, composite scores on the NIHTB-CB were moderately correlated with comparable composites from the WASI-II for both participants with CHD and typically developing peers, with no significant group differences. For the most part, subtests from the NIHTB-CB were significantly correlated with neuropsychological measures assessing similar constructs across groups, with small-to-medium effect sizes. That being said, the correlations between the NIHTB-CB DCCST and both the D-KEFS VFT Switching and the D-KEFS TMT Number Letter were not significant for either group, likely reflecting that the comparison measures do not sufficiently assess the same construct as the NIHTB-CB DCCST. Moreover, the positive correlation between the NIHTB-CB FIC+AT and the D-KEFS CWIT Inhibition was only significant among participants with CHD; however, the difference between the groups was not statistically significant. The positive correlation between the NIHTB-CB Oral Reading Test and WASI-II Vocabulary was only significant among healthy controls, and this represented the only statistically significant difference between participants with CHD and healthy controls.

**Table 2.**
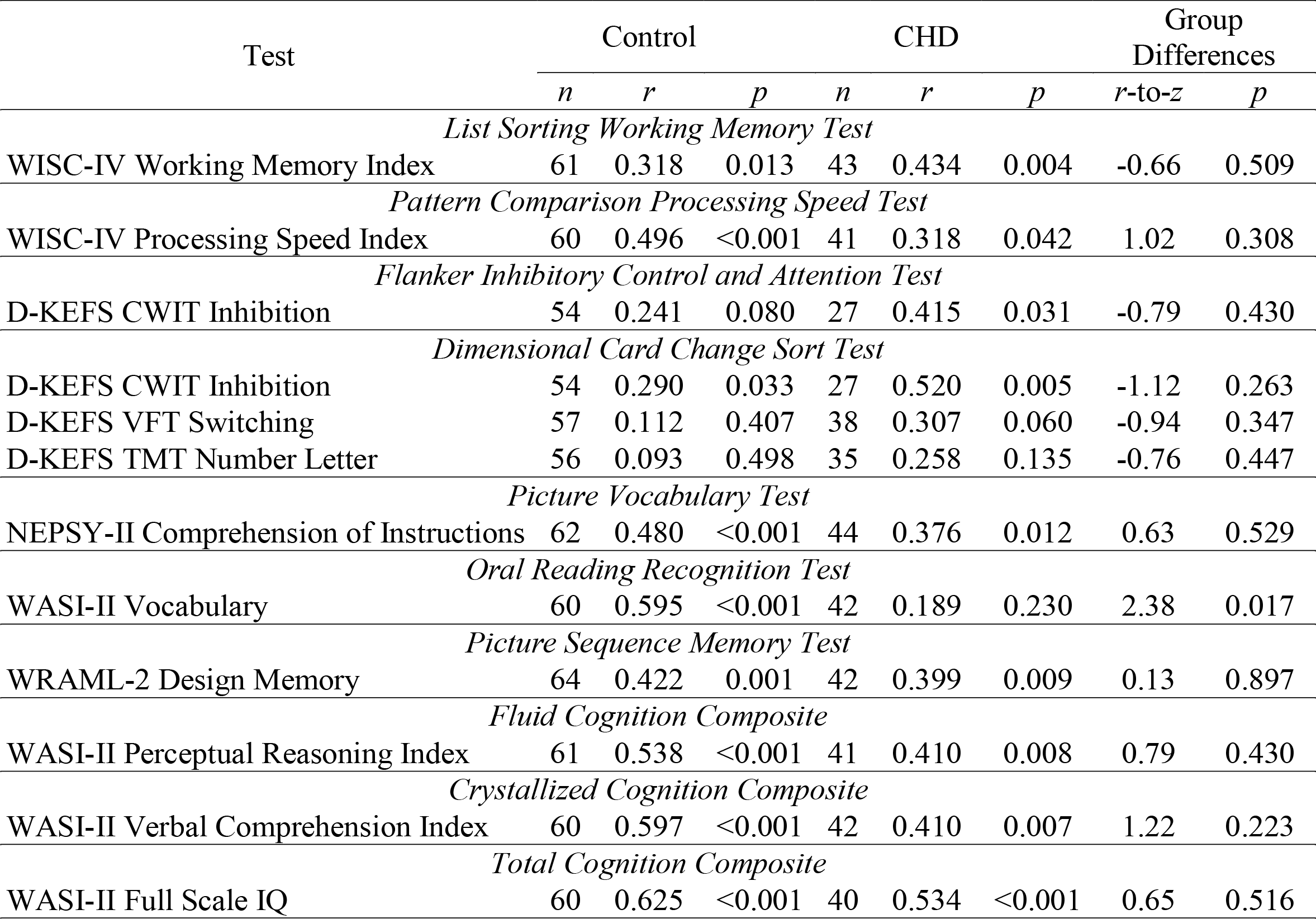
Convergent Validity Estimates Comparing the NIHTB-CB to Clinician-Issued Tests of Similar Domains

| Test | Control |  |  | CHD |  |  | Group Differences |  |
| --- | --- | --- | --- | --- | --- | --- | --- | --- |
|  | *n* | *r* | *p* | *n* | *r* | *p* | *r*-to-*z* | *p* |
| *List Sorting Working Memory Test* |  |  |  |  |  |  |  |  |
| WISC-IV Working Memory Index | 61 | 0.318 | 0.013 | 43 | 0.434 | 0.004 | -0.66 | 0.509 |
| *Pattern Comparison Processing Speed Test* |  |  |  |  |  |  |  |  |
| WISC-IV Processing Speed Index | 60 | 0.496 | <0.001 | 41 | 0.318 | 0.042 | 1.02 | 0.308 |
| *Flanker Inhibitory Control and Attention Test* |  |  |  |  |  |  |  |  |
| D-KEFS CWIT Inhibition | 54 | 0.241 | 0.080 | 27 | 0.415 | 0.031 | -0.79 | 0.430 |
| *Dimensional Card Change Sort Test* |  |  |  |  |  |  |  |  |
| D-KEFS CWIT Inhibition | 54 | 0.290 | 0.033 | 27 | 0.520 | 0.005 | -1.12 | 0.263 |
| D-KEFS VFT Switching | 57 | 0.112 | 0.407 | 38 | 0.307 | 0.060 | -0.94 | 0.347 |
| D-KEFS TMT Number Letter | 56 | 0.093 | 0.498 | 35 | 0.258 | 0.135 | -0.76 | 0.447 |
| *Picture Vocabulary Test* |  |  |  |  |  |  |  |  |
| NEPSY-II Comprehension of Instructions | 62 | 0.480 | <0.001 | 44 | 0.376 | 0.012 | 0.63 | 0.529 |
| *Oral Reading Recognition Test* |  |  |  |  |  |  |  |  |
| WASI-II Vocabulary | 60 | 0.595 | <0.001 | 42 | 0.189 | 0.230 | 2.38 | 0.017 |
| *Picture Sequence Memory Test* |  |  |  |  |  |  |  |  |
| WRAML-2 Design Memory | 64 | 0.422 | 0.001 | 42 | 0.399 | 0.009 | 0.13 | 0.897 |
| *Fluid Cognition Composite* |  |  |  |  |  |  |  |  |
| WASI-II Perceptual Reasoning Index | 61 | 0.538 | <0.001 | 41 | 0.410 | 0.008 | 0.79 | 0.430 |
| *Crystallized Cognition Composite* |  |  |  |  |  |  |  |  |
| WASI-II Verbal Comprehension Index | 60 | 0.597 | <0.001 | 42 | 0.410 | 0.007 | 1.22 | 0.223 |
| *Total Cognition Composite* |  |  |  |  |  |  |  |  |
| WASI-II Full Scale IQ | 60 | 0.625 | <0.001 | 40 | 0.534 | <0.001 | 0.65 | 0.516 |

#### Concurrent Validity

When considering whether scores from the NIHTB-CB were associated with measures of day-to-day functioning in a predictable fashion (i.e., assessing concurrent validity), we limited our analyses to composite scores of the NIHTB-CB. As shown in Table 3, for children and adolescents with CHD, better fluid cognition on the NIHTB-CB was associated with fewer externalizing and behavior problems, as rated by parents on the BASC-2. Better crystalized cognition on the NIHTB-CB was associated with better adaptive functioning across composite scores from the ABAS-II and BASC-2 as well as fewer behavior problems as indexed on the BASC-2. Better overall cognition on the NIHTB-CB was associated with better global and practice adaptive functioning on the ABAS-II and BASC-2 as well as fewer internalizing, externalizing, and behavior problems on the BASC-2. Although correlations were not significant for healthy control, there were no significant differences between participants with CHD and healthy controls for any of the analyses.

**Table 3.**
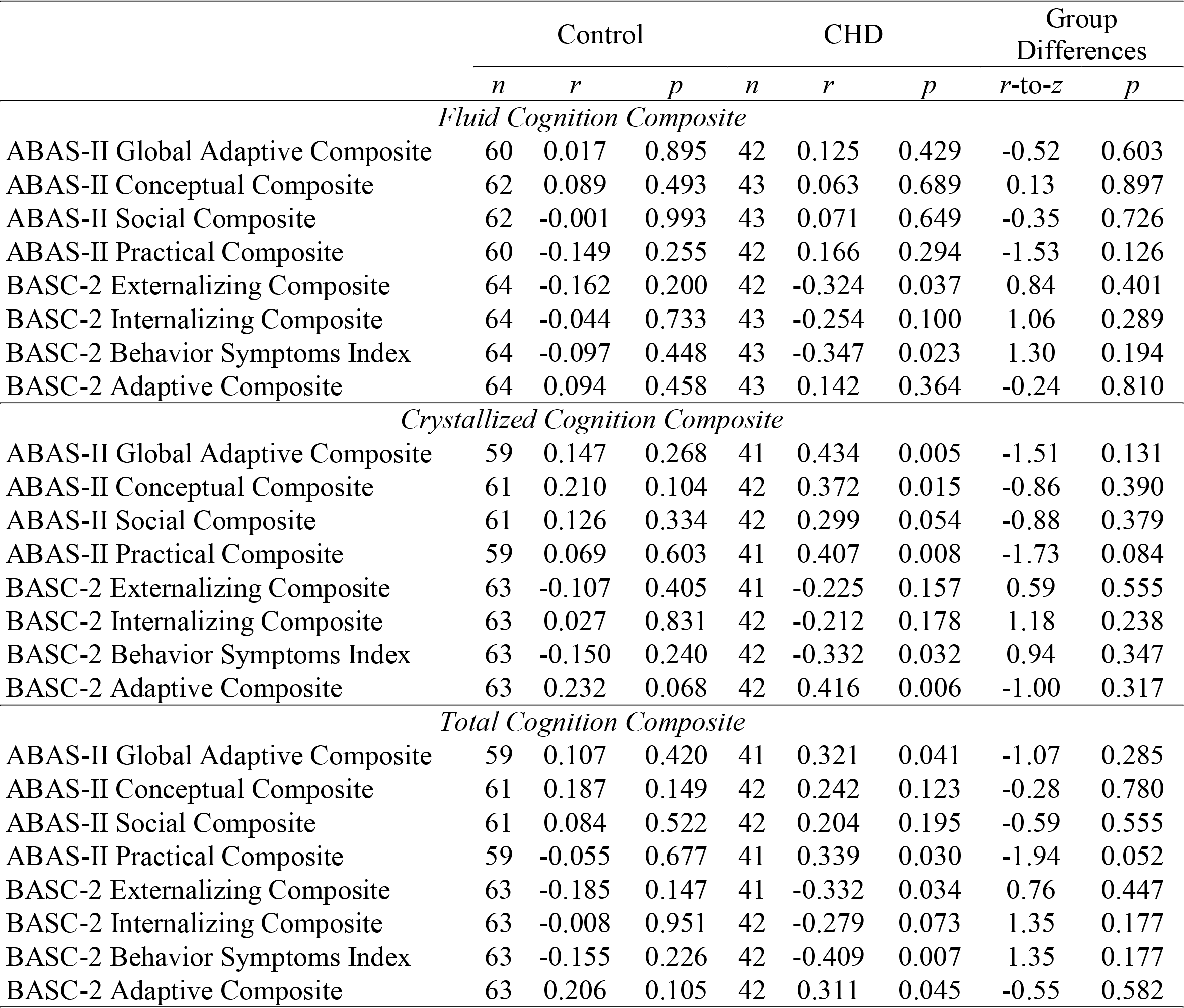
Concurrent Validity Estimates Comparing the NIHTB-CB to Clinician-Issued Ratings of Functioning

|  | Control |  |  | CHD |  |  | Group Differences |  |
| --- | --- | --- | --- | --- | --- | --- | --- | --- |
|  | *n* | *r* | *p* | *n* | *r* | *p* | *r-to-z* | *p* |
| *Fluid Cognition Composite* |  |  |  |  |  |  |  |  |
| ABAS-II Global Adaptive Composite | 60 | 0.017 | 0.895 | 42 | 0.125 | 0.429 | -0.52 | 0.603 |
| ABAS-II Conceptual Composite | 62 | 0.089 | 0.493 | 43 | 0.063 | 0.689 | 0.13 | 0.897 |
| ABAS-II Social Composite | 62 | -0.001 | 0.993 | 43 | 0.071 | 0.649 | -0.35 | 0.726 |
| ABAS-II Practical Composite | 60 | -0.149 | 0.255 | 42 | 0.166 | 0.294 | -1.53 | 0.126 |
| BASC-2 Externalizing Composite | 64 | -0.162 | 0.200 | 42 | -0.324 | 0.037 | 0.84 | 0.401 |
| BASC-2 Internalizing Composite | 64 | -0.044 | 0.733 | 43 | -0.254 | 0.100 | 1.06 | 0.289 |
| BASC-2 Behavior Symptoms Index | 64 | -0.097 | 0.448 | 43 | -0.347 | 0.023 | 1.30 | 0.194 |
| BASC-2 Adaptive Composite | 64 | 0.094 | 0.458 | 43 | 0.142 | 0.364 | -0.24 | 0.810 |
| *Crystallized Cognition Composite* |  |  |  |  |  |  |  |  |
| ABAS-II Global Adaptive Composite | 59 | 0.147 | 0.268 | 41 | 0.434 | 0.005 | -1.51 | 0.131 |
| ABAS-II Conceptual Composite | 61 | 0.210 | 0.104 | 42 | 0.372 | 0.015 | -0.86 | 0.390 |
| ABAS-II Social Composite | 61 | 0.126 | 0.334 | 42 | 0.299 | 0.054 | -0.88 | 0.379 |
| ABAS-II Practical Composite | 59 | 0.069 | 0.603 | 41 | 0.407 | 0.008 | -1.73 | 0.084 |
| BASC-2 Externalizing Composite | 63 | -0.107 | 0.405 | 41 | -0.225 | 0.157 | 0.59 | 0.555 |
| BASC-2 Internalizing Composite | 63 | 0.027 | 0.831 | 42 | -0.212 | 0.178 | 1.18 | 0.238 |
| BASC-2 Behavior Symptoms Index | 63 | -0.150 | 0.240 | 42 | -0.332 | 0.032 | 0.94 | 0.347 |
| BASC-2 Adaptive Composite | 63 | 0.232 | 0.068 | 42 | 0.416 | 0.006 | -1.00 | 0.317 |
| *Total Cognition Composite* |  |  |  |  |  |  |  |  |
| ABAS-II Global Adaptive Composite | 59 | 0.107 | 0.420 | 41 | 0.321 | 0.041 | -1.07 | 0.285 |
| ABAS-II Conceptual Composite | 61 | 0.187 | 0.149 | 42 | 0.242 | 0.123 | -0.28 | 0.780 |
| ABAS-II Social Composite | 61 | 0.084 | 0.522 | 42 | 0.204 | 0.195 | -0.59 | 0.555 |
| ABAS-II Practical Composite | 59 | -0.055 | 0.677 | 41 | 0.339 | 0.030 | -1.94 | 0.052 |
| BASC-2 Externalizing Composite | 63 | -0.185 | 0.147 | 41 | -0.332 | 0.034 | 0.76 | 0.447 |
| BASC-2 Internalizing Composite | 63 | -0.008 | 0.951 | 42 | -0.279 | 0.073 | 1.35 | 0.177 |
| BASC-2 Behavior Symptoms Index | 63 | -0.155 | 0.226 | 42 | -0.409 | 0.007 | 1.35 | 0.177 |
| BASC-2 Adaptive Composite | 63 | 0.206 | 0.105 | 42 | 0.311 | 0.045 | -0.55 | 0.582 |

### Emotion Battery

#### Group Differences

As depicted in Table 1, there were no significant differences between participants with CHD and typically developing peers for any of the scales from the NIHTH-EB. Overall, both groups of children and adolescents endorsed generally healthy socioemotional functioning.

#### Confirmatory Factor Analysis

As detailed in Table 4, there was adequate construct coherence for questionnaires from the NIHTB-EB among healthy control, based on the SRMR and CFI from CFA models, echoing studies on the development of the tools (Salsman et al., 2013). Still, the SRMR and CFI only trended towards guidelines of acceptable fit for certain scales (i.e., Sadness, Positive Affect for Ages 8-12, and Perceived Stress for Ages 13-17) and suggested poor fit for one scale (i.e., Positive Affect for Ages 13-17). By comparison, for participants with CHD, the majority of scales did not yield CFA models with SRMR and CFI within guidelines. Even if fit indices often trended towards acceptable ranges, they did not for Positive Affect for Ages 8-12 and for Ages 13-17 and Self-Efficacy for Ages 8-12 and for Ages 13-17. Of note, CFA models across the groups typically yielded RMSEA suggestive of poor fit. However, our small sample sizes likely inflated the RMSEA, making it a less useful measures in evaluating model fit (Kenny, Kaniskan, & McCoach, 2014).

**Table 4.** Indices of Internal Consistency and Confirmatory Factor Analysis for the NIHTB-EB

| Scale |  |  | Control |  |  | CHD |  |  |  |  |
| --- | --- | --- | --- | --- | --- | --- | --- | --- | --- | --- |
| Fixed Forms | *n* | $\alpha$ | SRMR | RMSEA | CFI | *n* | $\alpha$ | SRMR | RMSEA | CFI |
| Anger | 68 | 0.876 | 0.041 | 0.141 | 0.959 | 52 | 0.770 | 0.063 | 0.169 | 0.901 |
| Emotional Support | 68 | 0.913 | 0.047 | 0.145 | 0.939 | 52 | 0.874 | 0.107 | 0.234 | 0.796 |
| Fear | 68<sup>a</sup> | 0.921 | 0.074 | 0.185 | 0.902 | 52<sup>a</sup> | 0.955 | 0.065 | 0.216 | 0.873 |
| Friendship | 68 | 0.822 | 0.075 | 0.202 | 0.895 | 52 | 0.883 | 0.043 | 0.140 | 0.963 |
| Loneliness | 68 | 0.895 | 0.048 | 0.151 | 0.923 | 52 | 0.841 | 0.138 | 0.258 | 0.728 |
| Perceived Hostility | 68 | 0.873 | 0.061 | 0.202 | 0.920 | 51 | 0.869 | 0.072 | 0.280 | 0.862 |
| Perceived Rejection | 68 | 0.896 | 0.049 | 0.216 | 0.937 | 51 | 0.833 | 0.057 | 0.147 | 0.944 |
| Sadness | 68<sup>a</sup> | 0.881 | 0.073 | 0.262 | 0.824 | 50<sup>a</sup> | 0.924 | 0.111 | 0.248 | 0.811 |
| Life Satisfaction 8-12 | 33 | 0.771 | 0.080 | 0.139 | 0.930 | 25 | 0.779 | 0.113 | 0.256 | 0.785 |
| Positive Affect 8-12 | 28 | 0.817 | 0.122 | 0.203 | 0.660 | 17 | 0.713 | 0.146 | 0.204 | 0.429 |
| Perceived Stress 13-17 | 34 | 0.606 | 0.174 | 0.229 | 0.577 | 26 | 0.797 | 0.100 | 0.023 | 0.992 |
| CAT Forms | *n* | Marginal Reliability | SRMR | RMSEA | CFI | *n* | Marginal Reliability | SRMR | RMSEA | CFI |
| Self-Efficacy 8-12 | 29 | 0.918 | 0.072 | 0.089 | 0.951 | 22 | 0.876 | 0.210 | 0.243 | 0.366 |
| Life Satisfaction 13-17 | 35 | 0.921 | 0.101 | 0.165 | 0.792 | 27 | 0.869 | 0.106 | 0.148 | 0.752 |
| Self-Efficacy 13-17 | 35 | 0.927 | 0.080 | 0.153 | 0.869 | 27 | 0.842 | 0.130 | 0.167 | 0.673 |
| Positive Affect 13-17 | 35 | -- | 0.243 | 0.403 | 0.445 | 27 | -- | 0.147 | 0.433 | 0.453 |

*Notes.* <sup>a</sup> denotes that a smaller *n* was used to calculate Cronbach's $\alpha$ due to differences in items completed across participants. The marginal reliability for Positive Affect 13-17 could not be computed in a reliable fashion either within or across groups due to the number of items that were completed by a small number of participants.

#### Internal Consistency

As shown in Table 4, analyses using Cronbach’s α revealed acceptable internal consistency between fixed form items among participants with CHD (α ≥ 0.713), similar to healthy controls (α ≥ 0.771), with one exception. Among healthy controls, but not those with CHD, Perceived Stress for Ages 13-17 demonstrated questionable internal consistency (α = 0.606). Similarly, for CAT forms, marginal reliability coefficients from IRT models were suggestive of acceptable internal consistency among both participants with CHD and healthy controls (≥ 0.842).

#### Convergent Validity

When considering the convergent validity of the self-completed questionnaires from the NIHTB-EB, we focused our analyses on self-reports from the PedsQL. Although research suggests that parents can view their children’s functioning in a divergent manner than children do (Jackson et al., 2015), we also considered parent-reports from the PedsQL, given that we were limited in the number of participants who completed the rating scale. We conducted exploratory analyses using relevant subscales from the parent-completed ABAS-II and BASC-2. As a similar pattern of findings emerged as with the parent-completed PedsQL, we limit our discussion to the PedsQL.

As shown in Table 5, correlations between the scales of the NIHTB-EB and scores from the PedsQL were largely within expectable directions. It should be noted, though, that not all effects, even those with medium effect sizes were significant, as a function of sample size. Importantly, there were no significant group differences, with one exception. Healthy controls who endorsed greater perceived rejection on the NIHTB-EB were rated by their parents as having poorer social functioning on the PedsQL, but the same was not true for children and adolescents with CHD.

**Table 5.** Convergent Validity Estimates Comparing the NIHTB-EB to Rating Scales of Similar Domains

| Scale / Test | Control |  |  | CHD |  |  | Group Differences |  |
| --- | --- | --- | --- | --- | --- | --- | --- | --- |
|  | *n* | *r* | *p* | *n* | *r* | *p* | *r-to-z* | *p* |
| *Anger – Ages 8-17* |  |  |  |  |  |  |  |  |
| PedsQL Parent Emotional Functioning | 58 | -0.146 | 0.274 | 34 | 0.010 | 0.957 | -0.70 | 0.484 |
| PedsQL Child Emotional Functioning | 39 | -0.738 | <0.001 | 14 | -0.628 | 0.016 | -0.60 | 0.549 |
| *Emotional Support – Ages 8-17* |  |  |  |  |  |  |  |  |
| PedsQL Parent Social Functioning | 58 | 0.331 | 0.011 | 34 | -0.086 | 0.630 | 1.92 | 0.055 |
| PedsQL Child Social Functioning | 39 | 0.159 | 0.334 | 14 | 0.209 | 0.473 | -0.15 | 0.881 |
| *Fear – Ages 8-17* |  |  |  |  |  |  |  |  |
| PedsQL Parent Emotional Functioning | 57 | -0.328 | 0.013 | 34 | -0.087 | 0.625 | -1.12 | 0.263 |
| PedsQL Child Emotional Functioning | 38 | -0.765 | <0.001 | 14 | -0.806 | <0.001 | 0.32 | 0.749 |
| *Friendship – Ages 8-17* |  |  |  |  |  |  |  |  |
| PedsQL Parent Social Functioning | 57 | 0.358 | 0.006 | 34 | 0.146 | 0.409 | 1.01 | 0.313 |
| PedsQL Child Social Functioning | 38 | 0.173 | 0.298 | 13 | 0.243 | 0.402 | -0.20 | 0.842 |
| *Loneliness – Ages 8-17* |  |  |  |  |  |  |  |  |
| PedsQL Parent Social Functioning | 57 | -0.432 | 0.001 | 34 | -0.302 | 0.083 | -0.67 | 0.503 |
| PedsQL Child Social Functioning | 38 | -0.286 | 0.082 | 14 | -0.444 | 0.111 | 0.53 | 0.596 |
| *Perceived Hostility – Ages 8-17* |  |  |  |  |  |  |  |  |
| PedsQL Parent Social Functioning | 58 | -0.237 | 0.074 | 34 | -0.031 | 0.861 | -0.94 | 0.347 |
| PedsQL Child Social Functioning | 39 | -0.417 | 0.008 | 14 | -0.412 | 0.144 | -0.02 | 0.984 |
| *Perceived Rejection – Ages 8-17* |  |  |  |  |  |  |  |  |
| PedsQL Parent Social Functioning | 58 | -0.430 | 0.001 | 33 | 0.069 | 0.705 | -2.33 | 0.020 |
| PedsQL Child Social Functioning | 39 | -0.396 | 0.013 | 14 | -0.334 | 0.243 | -0.21 | 0.834 |
| *Sadness – Ages 8-17* |  |  |  |  |  |  |  |  |
| PedsQL Parent Emotional Functioning | 58 | -0.347 | 0.008 | 33 | -0.069 | 0.702 | -1.29 | 0.197 |
| PedsQL Child Emotional Functioning | 39 | -0.602 | <0.001 | 14 | -0.514 | 0.060 | -0.37 | 0.711 |
| *Perceived Stress – Ages 13-17* |  |  |  |  |  |  |  |  |
| PedsQL Parent Emotional Functioning | 28 | -0.318 | 0.099 | 14 | -0.292 | 0.311 | -0.08 | 0.936 |
| PedsQL Child Emotional Functioning | 20 | -0.560 | 0.010 | 5 | -0.878 | 0.050 | 0.98 | 0.327 |
| *Life Satisfaction – Ages 8-17* |  |  |  |  |  |  |  |  |
| PedsQL Parent Total Functioning | 57 | 0.307 | 0.020 | 34 | 0.361 | 0.036 | -0.27 | 0.787 |
| PedsQL Child Total Functioning | 38 | 0.352 | 0.030 | 14 | 0.399 | 0.158 | -0.16 | 0.873 |
| *Positive Affect – Ages 8-17* |  |  |  |  |  |  |  |  |
| PedsQL Parent Emotional Functioning | 51 | 0.014 | 0.924 | 31 | 0.186 | 0.318 | -0.73 | 0.465 |
| PedsQL Child Emotional Functioning | 39 | 0.107 | 0.518 | 14 | 0.447 | 0.109 | -1.08 | 0.280 |
| *Self-Efficacy – Ages 8-17* |  |  |  |  |  |  |  |  |
| PedsQL Parent Psychosocial Functioning | 58 | 0.091 | 0.496 | 33 | 0.119 | 0.511 | -0.12 | 0.905 |
| PedsQL Child Psychosocial Functioning | 39 | 0.337 | 0.036 | 14 | 0.032 | 0.914 | 0.93 | 0.352 |

## Discussion

The current study examined the psychometric properties NIHTB-CB and NIHTB-EB among children and adolescents with CHD, as compared to healthy controls. Prior research on the development of the NIHTB-CB composite scores and underlying subtests has demonstrated adequate factor structure (CFI ≥ 0.725, TLI ≥ 0.689, RSMEA ≤ 0.059, SRMR ≤ 0.039), robust developmental effects across childhood (*r* ≥ 0.77), and expectable correlations with measures of similar constructs (*r* ≥ 0.34) among children and adolescents (Bauer & Zelazo, 2013; Mungas et al., 2013). Similarly, pediatric self-reports from the NIHTB-EB have been found to assess subdomains of psychosocial functioning in a coherent manner (using confirmatory factor analysis, CFI ≥ 0.913; RSMEA <u>≥</u> 0.057), include items that reliably measure the same constructs (Cronbach’s α ≥ 0.86), and relate to comparable measures in a predictable way (|*r*| ≥ 0.28) (Salsman et al., 2013), with one exception (i.e., Perceived Rejection for Ages 13–17).

It is important to note, though, that the psychometric properties of the NIHTB were established among typically developing individuals but may differ in clinical populations. Recently, researchers have begun to use the NIHTB among individuals with CHD, a population with an increased risk of deficits across multiple cognitive and socioemotional domains (Bellinger & Newburger, 2013). For example, the NIHTB-CB has been used to assess the efficacy of cognitive training interventions among children with hypoplastic left heart syndrome and multiple forms of critical CHD (Calderon et al., 2019; Siciliano et al., 2020). Meanwhile, researchers have used a virtual administration of NIHTB-EB to assess the stress felt by patients with CHD during the COVID-19 pandemic (Cousino et al., 2020). Although the NIHTB has already begun to be administered patients with CHD, the psychometric properties of the tool within this population have largely remained unexplored. Siciliano and colleagues (2020) did note that children with hypoplastic left heart syndrome (HLHS) performed more poorly than healthy controls on the fluid cognition composite score from the NIHTB-CB, and the fluid cognition composite score was positively associated, with medium to large effect sizes, with the fluid reasoning, working memory, and processing speed indices of the Wechsler Intelligence Scale for Children, 5^th^ edition (WISC-V). That being said, the study, limited to those with HLHS, did not consider associations of the individual subtests, the crystalized cognition composite score, or the overall composite.

The results of the current study provide the first comprehensive assessment of the psychometric properties of the NIHTB-CB and NIHTB-EB among children with CHD. As has been found in prior work with typically developing children, we found developmentally expected gains in performance across cognitive tasks for those with CHD. Although subjects with CHD largely performed within expectation for their age on cognitive tasks, there were inefficiencies among children and adolescents with CHD as compared to healthy controls on all cognitive tasks with exception of the LSWMT. Such a pattern of findings is consistent with prior research showing subtle cognitive vulnerabilities among children with CHD when considered as a whole (Bellinger & Newburger, 2013; Marino et al., 2012). For the most part, composites and subtests from the NIHTB-CB were significantly correlated with neuropsychological measures assessing similar constructs across groups, with small-to-medium effect sizes. Moreover, higher scores on the NIHTB-CB were also generally associated with ratings of better day-to-day functioning among children with CHD. As such, the brief and user-friendly NIHTB-CB appears to provide a valid and useful assessment of cognitive functioning among children and adolescents with CHD.

On the NIHTB-EB, we found that both children and adolescents with CHD and their typically developing peers endorsed generally healthy socioemotional functioning, and there were no significant differences between the groups for any of the scales. Prior research has similarly found that, although parents and other informants report socioemotional vulnerabilities, children with CHD often do not (Jackson et al., 2015). For the most part, scales from the NIHTB-EB showed acceptable internal consistency among both participants with CHD and healthy controls. Interestingly, there was adequate construct coherence for most of questionnaires from the NIHTB-EB among healthy control, echoing studies on the development of the tools (Salsman et al., 2013). However, for participants with CHD, the majority of scales did not yield models with adequate fit.

Correlations between the scales of the NIHTB-EB and a comparison tool were largely within expectable directions, although analyses were limited by our sample size. Overall, while the NIHTB-EB may be a reliable and valid tool among patients with CHD, further research is needed to understand the meaningfulness of the scales for participants with CHD.

Similar to our work with children and adolescents with CHD, there are emerging efforts to assess the feasibility and properties of the NIHTB among diverse groups. Among children and adolescents, the literature has, for example, supported the use of the cognitive battery among those with intellectual disability, traumatic and other acquired brain injuries, and epilepsy (Chadwick et al., 2021; Shields et al., 2020; Thompson et al., 2020; Watson et al., 2020). Still, research has found that the cognitive battery from the NIHTB may not be appropriate or sensitive to differences in functioning among those with a high degree of impairment or agitation. It has been noted that young children with intellectual disability and older children with very low functioning may require test adaptations (Shields et al., 2020). In fact, children and adolescents with a high degree of cognitive impairment may not be able to complete the NIHTB-CB, as noted in a study with pediatric patients with treatment-resistant epilepsy (Thompson et al., 2020). Similar findings have been seen with adult samples. Although the NIHTB-CB has been found to be a useful battery when considering Alzheimer’s disease, the tests of memory may be too difficult and insufficiently sensitive for those at the lower end of memory function (Hackett et al., 2018; Ma et al., 2021). It has also been noted that children and adolescents with a high degree agitation (e.g., inhibition, emotional lability, and aggression) may not be able to complete the NIHTB-CB, as noted in a study with pediatric patients with acquired brain injuries (Watson et al., 2020). Such findings across clinical populations point to limitations of the NIHTB-CB that may also be applicable to patients with CHD. Although, on average, those with CHD display intellectual functioning in the average range and no more than mild behavioral dysregulation, there are subtests of patients with cognitive impairments and agitation who may not be well-served by the NIHTB-CB. Indeed, in our study, there were several individuals who could not complete testing due to behavioral noncompliance.

Emerging work with other clinical populations has also found that the cognitive tests of the NIHTB may uniquely capture the subtle deficits seen in some groups. For instance, Chadwick and colleagues (2021) noted that the NIHTB-CB may be more advantageous for detecting cognitive deficits after mild TBI in pediatric patients compared to traditional tests given its focus on reaction time as well as accuracy when assessing attention and executive functioning. Yet, research has suggested that the brief cognitive battery from the NIHTB may be less sensitive in capturing subtle cognitive differences in other populations. For example, Meredith and colleagues (2020) did not find differences in cognition between adults with alcohol use disorder and healthy controls using the NIHTB-CB, as found in studies with comprehensive neuropsychological batteries. Data from the present study suggested that the NIHTB-CB is generally sensitive to differences in cognition between patients with CHD and healthy controls. Still, its measure of working memory did not pick up on the differences between the groups that were seen within traditional measures.

It is important to acknowledge, that while our study is the first to provide a comprehensive view of the psychometric properties of the NIHTB-CB and NIHTB-EB among children and adolescents with CHD, there are limitations to our findings. First, longitudinal data were not available in order to explore test-retest reliability, the ability to detect meaningful change, and predictive relationships. However, data among typically developing children and adolescents suggests that further longitudinal studies may be beneficial. Although prior work has found strong test-retest reliability at short intervals for the NIHTB-CB (*ICC* ≥ 0.76), moderate associations have been seen at multi-year intervals (ICC = 0.31–0.76) (Bauer & Zelazo, 2013; Taylor et al., 2020). Second, we were limited to the tools used within our larger study when considering how traditional measures compared to the NIHTB. Third, we acknowledge that we had a small sample size for certain analyses. As a result of the available tools and sample size, we were restricted in exploring the validity of the NIHTB-CB and, particularly, the NIHTB-EB, and it would be helpful to continue exploring this area in future research. Future studies may also investigate the effect of CHD lesion type and severity, social economic status, and other medical and demographic factors on the psychometric properties of the NIHTB (Loccoh et al., 2018; Marino et al., 2012; Naef at al., 2017).

Overall, the NIH Toolbox offers developmentally sensitive, reliable, and valid assessments of cognitive abilities and psychosocial functioning among children and adolescents with CHD. As such, the easy-to-administer, time-efficient, and cost-effective tool may facilitate clinical and empirical endeavors requiring brief and repeatable assessments. Still, the NIH Toolbox may not provide the breadth, level of detail, and sensitivity needed for all contexts. As such, clinicians and researchers may continue to need complementary measures for comprehensive assessments.

## Data Availability

All data produced in the present study are available upon reasonable request to the author.

## Abbreviations

ABAS-II: Adaptive Behavior Assessment System, 2^nd^ edition
BASC-2: Behavior Assessment System for Children, 2^nd^ edition
CAT: computer-adaptive testing
CFA: confirmatory factor analyses
CFI: comparative fit index
CHD: congenital heart defects
CWIT: Color-Word Interference Test
DCCST: Dimensional Change Card Sort Test
D-KEFS: Delis-Kaplan Executive Function Scale
FIC+AT: Flanker Inhibitory Control and Attention Test
HLHS: hypoplastic left heart syndrome
IRT: item response theory
MICE: multiple imputation by chained equations
NEPSY-2: Neuropsychology Assessment, 2^nd^ edition
NIHTB: National Institutes of Health Toolbox
NIHTB-CB: NIHTB Cognition Battery
NIHTB-EB: NIHTB Emotion Battery
LSWMT: List Sorting Working Memory Test
ORRT: Oral Reading Recognition Test
PCPST: Pattern Comparison Processing Speed Test
PedsQL: Pediatric Quality of Life Inventory
PSMT: Picture Sequence Memory Test
PVT: Picture Vocabulary Test
RMSEA: root mean square error of approximation
SRMR: standardized root mean square residual
TMT: Trail Making Test
VFT: Verbal Fluency Test
WASI-II: Wechsler Abbreviated Scale of Intelligence, 2^nd^ edition
WISC-IV: Wechsler Intelligence Scale for Children, 4^th^ edition
WISC-V: Wechsler Intelligence Scale for Children, 5^th^ edition
WRAML-2: Wide Range Assessment of Memory and Learning, 2^nd^ edition

---

**Supplemental Table 1.** Demographics Split by Group and Distribution of Congenital Heart Defects and Intracranial Injury / Abnormalities Among Participants with CHD

| Variable / Diagnosis | Control | CHD | $X^2 / t$ |
| --- | --- | --- | --- |
|  | *n* (%) | *n* (%) | *p* |
| Males (%) | 43 (53.75%) | 45 (77.59%) | 0.0040 |
| White (%) | 52 (65.00%) | 50 (86.21%) | 0.0051 |
| Right-Hand Dominant (%) | 65 (87.84%) | 47 (81.03%) | 0.2793 |
| Preterm Birth (%) | 8 (13.11%) | 8 (14.04%) | 0.8840 |
| Mean Maternal Education (*SD*) | 1.78 (0.52) | 1.61 (0.49) | 0.0563 |
| Mean Household Income (*SD*) | 3.65 (2.13) | 3.39 (1.83) | 0.5303 |
| Mean Age at Assessment (*SD*) | 12.63 (2.96) | 12.46 (3.35) | 0.7604 |
| Congenital Heart Defect (CHD) |  |  |  |
| Single Ventricle (SV) | - | 26 (44.83%) | - |
| Hypoplastic Left Heart Syndrome (HLHS) | - | 13 (22.41%) | - |
| Double Inlet Left Ventricle (DILV) | - | 5 (8.62%) | - |
| Double Outlet Right Ventricle (DORV) | - | 9 (15.52%) | - |
| Aortic Arch Anomaly | - | 38 (65.52%) | - |
| Coarctation of Aorta (COA) | - | 8 (13.79%) | - |
| Hypoplastic / Interrupted Aortic Arch (HAA / IAA) | - | 4 (6.90%) | - |
| Bicuspid Aortic Arch (BAV) | - | 10 (17.24%) | - |
| Aortic Valve / Pulmonary Stenosis (AVS / PS) | - | 23 (39.66%) | - |
| Total / Partial Anomalous Pulmonary Venous Return (TAPVR / PAPVR) | - | 3 (5.17%) | - |
| Tetralogy of Fallot (TOF) | - | 4 (6.90%) | - |
| Transposition of Great Arteries (TGA) | - | 13 (22.41%) | - |
| Atrioventricular Septal Defect (AVSD) | - | 3 (5.17%) | - |
| Ventricular / Atrial Septal Defect (VSD / ASD) | - | 28 (48.28%) | - |
| Cardiac Surgery | - | 52 (89.66%) | - |
| Age at 1<sup>st</sup> Cardiac Surgery ≤ 1 Year | - | 39 (67.24%) | - |
| Age at 1<sup>st</sup> Cardiac Surgery ≤ 7 Days | - | 19 (32.76%) | - |
| Intracranial Injury / Abnormality |  |  |  |
| Hemorrhage | - | 4 (6.90%) | - |
| Stroke | - | 6 (10.34%) | - |
| Encephalomalacia | - | 2 (3.45%) | - |
| Ventriculomegaly / Hydrocephalus | - | 2 (3.45%) | - |
| Gliosis | - | 3 (5.17%) | - |
| Necrosis | - | 1 (1.72%) | - |
| Volume Loss | - | 5 (8.62%) | - |
| Structural Abnormalities | - | 6 (10.34%) | - |

*Note.* Maternal Education: 0 = no high school diploma, 1 = high school diploma, 2 = college graduate and above. Household Income: 0 = less than \$25,000, 1 = \$25,000 to \$34,999, 2 = \$35,000 to \$49,999, 3 = \$50,000 to \$74,999, 4 = \$75,000 to \$99,999, 5 = \$100,000 to \$149,999; 6 = \$150,000 or more.

**Supplemental Table 2.** Group Differences for Clinician-Issued Battery and Rating Scales

| Test / Composite / Scale | Control |  | CHD |  | *t* | *p* |
| --- | --- | --- | --- | --- | --- | --- |
| | *n* | $\mu$ (*SD*) | *n* | $\mu$ (*SD*) | | |
| WASI-II: Vocabulary | 61 | 57.61 (9.768) | 44 | 53.98 (8.851) | 1.953 | 0.054 |
| Verbal Comprehension Index | 61 | 110.70 (14.024) | 44 | 105.70 (12.275) | 1.898 | 0.061 |
| Perceptual Reasoning Index | 61 | 108.41 (15.174) | 42 | 99.52 (18.081) | 2.700 | 0.008 |
| Full Scale IQ | 61 | 110.67 (13.314) | 42 | 102.79 (15.889) | 2.729 | 0.008 |
| NEPSY-2: Comprehension of Instructions | 69 | 11.74 (2.984) | 44 | 10.41 (2.773) | 2.374 | 0.019 |
| WISC-IV: Digit Span | 67 | 10.45 (3.001) | 44 | 9.00 (2.745) | 2.570 | 0.012 |
| Letter Number Sequencing | 61 | 11.15 (2.072) | 44 | 9.50 (2.758) | 3.341 | 0.001 |
| Coding | 66 | 10.56 (3.379) | 42 | 8.26 (3.140) | 3.541 | 0.001 |
| Symbol Search | 66 | 11.74 (2.963) | 42 | 10.10 (2.621) | 2.943 | 0.004 |
| Working Memory Index | 67 | 103.75 (12.679) | 44 | 95.02 (13.498) | 3.456 | 0.001 |
| Processing Speed Index | 66 | 106.65 (16.051) | 42 | 95.45 (14.397) | 3.676 | <0.001 |
| D-KEFS: TMT – Number Letter Switching | 62 | 10.94 (2.268) | 35 | 8.31 (2.888) | 4.944 | <0.001 |
| VFT – Switching | 63 | 11.33 (3.188) | 38 | 10.32 (2.672) | 1.648 | 0.102 |
| CWIT – Inhibition | 54 | 10.80 (2.326) | 27 | 8.74 (2.982) | 3.406 | 0.001 |
| WRAML-2: Design Memory | 70 | 9.90 (2.959) | 42 | 7.64 (2.448) | 4.160 | <0.001 |
| ABAS-II: Global Adaptive Composite | 66 | 104.67 (13.324) | 43 | 95.26 (17.159) | 3.213 | 0.002 |
| Conceptual Composite | 68 | 106.71 (12.930) | 44 | 96.86 (14.767) | 3.719 | <0.001 |
| Social Composite | 68 | 105.46 (13.895) | 44 | 98.95 (15.622) | 2.302 | 0.023 |
| Practical Composite | 66 | 101.70 (13.791) | 43 | 92.23 (16.883) | 3.202 | 0.002 |
| BASC-2: Externalizing Composite | 70 | 43.73 (8.446) | 43 | 46.05 (8.588) | -1.407 | 0.162 |
| Internalizing Composite | 70 | 43.54 (9.234) | 44 | 53.00 (13.701) | -4.038 | <0.001 |
| Behavior Symptoms Index | 70 | 43.89 (7.700) | 44 | 48.75 (11.115) | -2.544 | 0.013 |
| Adaptive Composite | 70 | 55.50 (11.553) | 44 | 48.64 (12.529) | 2.989 | 0.003 |
| PedsQL: Parent Report – Total | 64 | 91.73 (9.623) | 44 | 71.737 (19.349) | 6.338 | <0.001 |
| Child Report – Total | 42 | 81.55 (13.497) | 16 | 72.92 (13.486) | 2.178 | 0.034 |

*Note.* WASI-II: *T*-scores for subtests ( $M = 50$ , $SD = 10$ ) and standard scores for composites ( $M = 100$ , $SD = 15$ ). D-KEFS: scaled scores ( $M = 10$ , $SD = 3$ ). NEPSY-2: scaled scores ( $M = 10$ , $SD = 3$ ). WISC-IV: scaled scores for subtests ( $M = 10$ , $SD = 3$ ) and standard scores for composites ( $M = 100$ , $SD = 15$ ).
WRAML-2: scaled score ( $M = 10$ , $SD = 3$ ). ABAS-II: standard scores ( $M = 100$ , $SD = 15$ ). BASC-2: *T*-scores ( $M = 50$ , $SD = 10$ ).

